# Mucinous Pancreatic Cysts: A Single-Center Experience Epidemiology, Surveillance, Risk Formulation, Surgery

**DOI:** 10.64898/2026.11.23.26368751

**Authors:** Rick Selby, Kurtis Liang, Ralitza Parina, Isabella Livits, Giselle Tran, Phuong Nguyen, Linda Sher, Paul Korc, Ravi Kankotia, Marcus Breit, David Braxton, Sophie Pénisson, Cristian Tomasetti, Daniel Von Hoff, Steven R. Grossman

**Affiliations:** Hoag Memorial Hospital Presbyterian, Newport Beach, CA; Keck Medicine of USC, Los Angeles, CA; Translational Genomics Research Institute, Phoenix, AZ

## Abstract

**Background:** Mucinous pancreatic cysts demonstrate malignant risk and expanding prevalence. Society guidelines lack underpinning from large, single-center experiences. We describe pancreatic mucinous cyst epidemiology, compare cyst-derived cancers from surveilled vs. non-surveilled cohorts, analyze risk metrics using surveillance and pathologic outcomes, and propose a weighted cancer risk probability score.

**Methods:** We conducted a retrospective study of patients at a tertiary metropolitan hospital between January 2006 and December 2025 who were found to have a mucinous pancreatic cyst or cyst-derived cancer on endoscopic ultrasound (EUS). A diagnostic algorithm was used to presumptively diagnose cyst phenotypes; diagnoses were clarified through surveillance and surgery. A preliminary risk classification scheme categorized patients to a risk level based on six binary variables. Cyst-derived cancer (CDCa) were categorized by first encounter or progression from surveillance. Survival analysis of surgical patients was organized by histologic grade and tumor stage. We developed a new malignant progression risk formula that weighted the four most predictive risk metrics: cyst size, main duct phenotype, pancreatic duct dilation, and bile duct dilation.

**Results:** 3667 cysts in 3643 patients were analyzed; 2087 (57%) were branch duct and 93 (2.9%) were main duct intraductal papillary mucinous neoplasms (IPMN), and 79 (2.4%) were mucinous cystic neoplasms (MCN). Unclassifiable cysts (n = 1386, 32%) were analyzed as a primordial branch duct IPMN cohort due to volume and gender equivalents. IPMN-MD were often mislabeled as IPMN-BD during surveillance (43%). Our initial recommended surveillance interval was based on six unweighted metrics: solid nodule, pancreatic or bile duct dilation, main duct involvement, and fluid chemistry.

Cyst-derived cancers developed in 8% of mucinous cysts (n = 162). Resection was performed in 184 patients from surveilled (n = 76) and non-surveilled (n = 108) cohorts. Surveilled patients had fewer advanced stage cancers; however, explant pathology was low/moderate dysplasia in 72 resected patients.

Kaplan-Meier survival at 5 years was 82% with high-grade dysplasia and T1N0 and 50% for T2N0 and T3N0 lesions. Frequencies of malignant cyst progression were main duct IPMNs (53%), branch duct IPMN (6%), and MCN (5%). Mucinous cyst-derived cancers accounted for 10.7% of all pancreatic adenocarcinomas within our registry.

Our logistic regression model achieved a cross-validated area under the ROC of 0.957 (95% CI: 0.944–0.968), with 90% sensitivity at a threshold corresponding to 90% specificity.

**Conclusion:** In this longitudinal cohort study, branch duct IPMN was the dominant phenotype, yet main duct IPMN was most likely to progress to malignancy. Surveillance favored early cancer diagnosis yet with low specificity. Post resection survival was excellent and explant pathology revealed lower frequency of nodal disease when compared to PDAC. Risk metric analysis led to a new risk-formulation strategy for patient selection. Our new weighted risk formula offers high predictive accuracy, helping to avoid unnecessary resections.

## Introduction

The prevalence of pancreatic cystic lesions (PCLs) nearly doubled from 2010 to 2017, an annual growth rate of nearly 9%.^1^ Most are asymptomatic and identified through cross-sectional imaging obtained for unrelated indications, with up to 3% of CT scans and 20% of MRIs demonstrating incidental PCLs.^2,3^

Of radiologically identified PCL’s, the vast majority (80%) are intraductal papillary mucinous neoplasms (IPMNs) which were only formally defined by the World Health Organization in 1996.^3,4^

The clinical importance of mucinous pancreatic cystic lesions stems from their malignant potential, which contributes to overall population pancreatic cancer incidence. Uncertainty remains regarding the rate of progression from cysts to cancer as well as the optimal surveillance strategy once identified. ^5,6^ Crippa et al. showed that within five years of follow-up, one in six patients with IPMNs develop worrisome features or high-risk stigmata indicating progression toward malignancy. ^7^ Long-term studies found that approximately 1 in 12 patients with IPMNs develop pancreatic cancer beyond 10 years of surveillance, emphasizing the need for both sensitive and specific long-term monitoring strategies. ^8,9^

Most studies examining PCL surveillance have been retrospective. Current prospective studies of presumed mucinous cyst phenotypes face limitations of small cohort sizes and selection bias. Inconsistent follow-up protocols have led to variable estimates of the risk of malignant progression.^1,10^ While mucinous cysts alone do not mandate resection, their prevalence and malignant predisposition justify structured surveillance with clearly defined operative triggers. Earlier management strategies were often indiscriminate and associated with surgical over-inclusion.^11,12^ Much of the current understanding of malignancy risk derives from surgical series, which estimate rates of high-grade dysplasia or invasive carcinoma of up to 70% in main-duct IPMNs (IPMN-MD) and 30% in branch-duct IPMNs (IPMN-BD).^3^

Risk-prediction frameworks, such as Sendai, AGA, Kyoto and European guidelines, rely predominantly on anatomic imaging features, which don’t reliably detect high-grade dysplasia or microinvasive cancer at what is perceived as a surgically curable stage.^8–10,12–17^ Endoscopic ultrasound (EUS) has expanded diagnostic capability through high-resolution imaging, targeted fine-needle aspiration, and cyst fluid analysis, improving differentiation of mucinous from non-mucinous lesions as well as earlier malignancy detection.^18,19^ Cyst fluid analysis also has the potential to improve previous guidelines.^20^

Incremental increases in mucinous PCL incidence have led to increased imaging of the population and resulted in large patient volumes requiring long-term surveillance, burdening programs with logistical complexities in the form of workflows, data tracking, and healthcare financing. ^21–24^Broad risk stratification based on perceived risk of malignant progression is needed to reduce unnecessary surveillance for low-risk individuals while intensifying observation and intervention for those at higher risk. Longitudinal data is important to determine surveillance endpoints.

Due to the increasing population health burden of PCL surveillance, large single-center or multi-institutional studies that incorporate surgical outcomes are essential to accurately determine the accuracy of pancreatic cyst surveillance algorithms and the natural history of the mucinous phenotypes. In this work, we describe the epidemiology, long-term surveillance, and surgical outcomes of a large, single-center PCL cohort, and validate a robust risk stratification model that can efficiently guide prospective management and surveillance of mucinous PCLs to identify patients at risk for progression to malignancy.

## Methods

This study received Institutional Review Board approval.

### Patient population

A single-center retrospective analysis of patients with mucinous pancreatic cystic lesions (M-PCLs) in a single institution was performed. M-PCL patients who underwent EUS at our hospital between January 2006 and December 2025 were included in the study. Patients fell broadly into three categories based on referral patterns, 1) discovery of an asymptomatic M-PCL, 2) symptomatic M-PCL management, and 3) an initial presentation diagnosed as mucinous cyst-derived cancer (CDCa). The first group was placed in surveillance while the latter two groups were managed operatively or referred to medical oncology.

### Data collection

All patients began surveillance with an endoscopic ultrasound (EUS) and data was recorded for each cyst identified. Cysts were classified according to a pre-determined cyst classification scheme, with each patient assigned a recommended surveillance interval based on a risk adjustment model described below. Total time in surveillance for M-PCL patients was measured as days between initial EUS and the most recent imaging study. Patients were deemed active in surveillance if they were compliant with their assigned follow-up.

Collected data included patient demographics, family history, anatomic variables (cyst location, pancreatic duct connection, solid component, pancreatic duct dilation, common bile duct dilation), cytologic data (mucin staining, cytology, cyst CEA level, cyst amylase level), and genetic data (risk category based on commercially available testing classification, DNA quantity and quality, KRAS mutation, GNAS mutation, loss of heterozygosity). Pathology data was obtained from surgical explants.

### M-PCL labeling

M-PCLs were classified presumptively based on a combination of imaging, cytologic studies, and biopsies where available. This “inferred” phenotypic diagnosis was progressively clarified as patients underwent subsequent imaging and cyst sampling. This allowed the dataset to serve as a dynamic and comprehensive account of M-PCLS throughout each patient’s surveillance. For patients who had multiple cysts, each cyst >9mm was tracked in the database separately over time. Cysts were labeled as “unclassifiable” if they had no diagnostic anatomic features and were too small to aspirate. However, such cysts were considered possible M-PCLs and surveilled expectantly. Non-mucinous cyst data was collected as part of the program but not included in the analytic portion of the study.

### Derivation of risk score and statistical analysis

Early in the study, an empiric risk classification scheme was used to assign patients a recommended surveillance imaging frequency. Patients were classified as low, intermediate, or high risk based on six factors: cyst size, presence of solid nodule, biliary or pancreatic ductal dilatation, presumed main duct IPMN, and molecular analysis, as described in **Table 1**. For patients with strong syndromic family cancer history or possible cyst-associated symptoms (pancreatic colic), these were considered as modifiers of the surveillance interval or surgical indication despite not strictly a part of the stratification scheme.

**Table 1:**
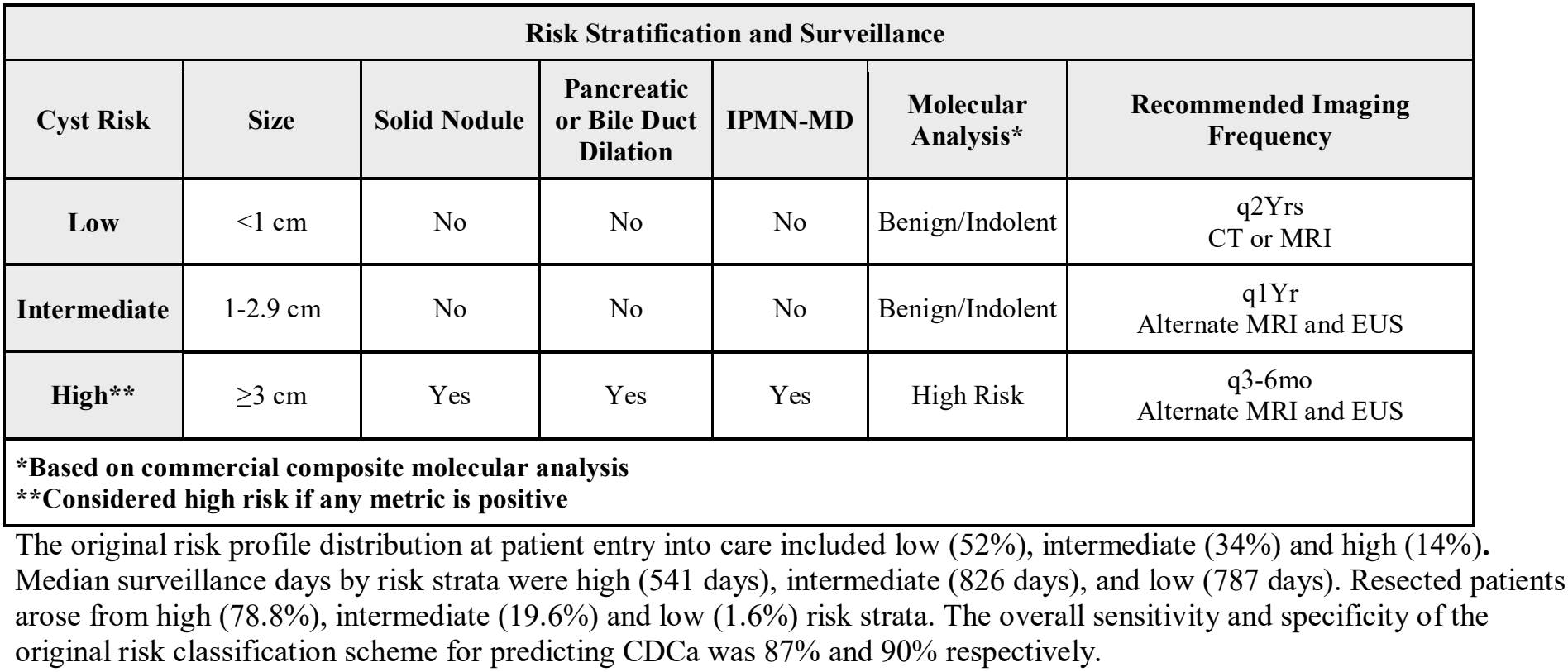
Original risk stratification scheme.

As more patient data was collected, a new, data-driven risk stratification score (RSS) was developed and internally validated. Two multivariable logistic regression models were constructed: a full model including all six predictors, and a reduced model including four predictors (cyst size, main-duct IPMN phenotype, pancreatic duct dilation, and bile duct dilation). Logistic regression was used as a statistical approach to combine these predictors into a single risk score. The risk score results in a value between 0 and 1, reflecting the relative predicted probability that a cyst is malignant. Model performance was evaluated using a 10-fold cross-validated area under the receiver operating characteristic (ROC) curve (AUC), a measure of how well the model distinguishes malignant from benign cysts. An AUC of 1 indicates perfect discrimination, whereas 0.5 indicates discrimination no better than chance. Risk scores were calculated for all patients, and the final model was selected based on discriminative ability and parsimony.

### Analysis of CDCa

Broadly, patients fell into two cohorts: those whose CDCa was present at initial presentation (***ip***CDCa) and those who entered care with a mucinous cyst of presumed phenotype and developed CDCa during surveillance (***sv***CDCa). Final surgical pathology was grouped in four categories: 1) benign lesions (including low and moderate-grade dysplasia); 2) early-stage CDCa (high-grade dysplasia, T1μ, T1N0), 3) mid-stage CDCa (T2N0, T3N0), and 4) late-stage CDCa (>T3 or >N0).

Descriptive statistics comparing the distribution of pathology between ***sv***CDCa and ***ip***CDCa were performed. Kaplan-Meier survival analysis was performed comparing both cohorts as well. SPSS Version 30 was used for statistical analysis and generation of Kaplan-Meier survival curves.

Using our institutional cancer registry data from 2006-2025, we identified all pancreatic ductal adenocarcinoma patients diagnosed in our center to calculate the contribution from CDCa.

### Surgical outcomes

Surgical resections for presumed CDCa were included in this portion of the analysis.

Procedure type, margin status, final pathology, surveillance status, and disease-free survival were collected.

### Analysis of surveillance failures

Surveillance fallouts that resulted in CDCa were scrutinized to determine if the risk stratification score failed due to:

#### Progression failure

Progression of a mucinous cyst beyond T1N0 malignancy in a patient who was compliant with surveillance. This suggests underperformance of the risk stratification scheme by allowing a tumor to progress, reducing curability while in surveillance.

#### Compliance disruption failure

Progression of a mucinous cyst to malignancy related to disruption of patient-program contact. This suggests both patient compliance failures as well as logistics and workflow failures on behalf of the program.

### Analysis of Tumor Generative Index (TGI)

TGI is the percentage of CDCa arising from a particular M-PCL phenotype from the total number of patients within that same phenotype.

### Patient inactivity

Given the natural history and chronicity of pancreatic cysts, patients may remain in surveillance for years. The database was regularly reconciled by analyzing patient programmatic activity and grouping reasons for patient inactivity. Inactive patient records were kept in the database for patient analytics and the possibility of returning to care. Patients were classified as “aged out” if they had a presumed short life expectancy or severe comorbidities. The remaining reasons for inactive designation are listed in **Supplemental Table 1**.

## Results

### Study population

We identified 4278 PCLs from 4179 patients. We excluded non-mucinous PCLs (n = 611, 14.3%) and analyzed the M-PCL (n = 2281, 53.3%), and unclassifiable PCL (n = 1386, 32.4%) cohorts together. The combined study cohort included 3667 PCLs. M-PCL phenotypes included IPMN-BD (n = 2087, 91.5%), IPMN-MD (n = 115, 5.0%) and mucinous cystic neoplasm (MCN) (n = 79, 3.5%). Unclassifiable cysts had parallel numbers, similar gender equivalents, and younger age compared to M-PCLs. **Table 2** gives demographics and features of M-PCL phenotypes and unclassifiable cyst cohorts.

**Table 2:**
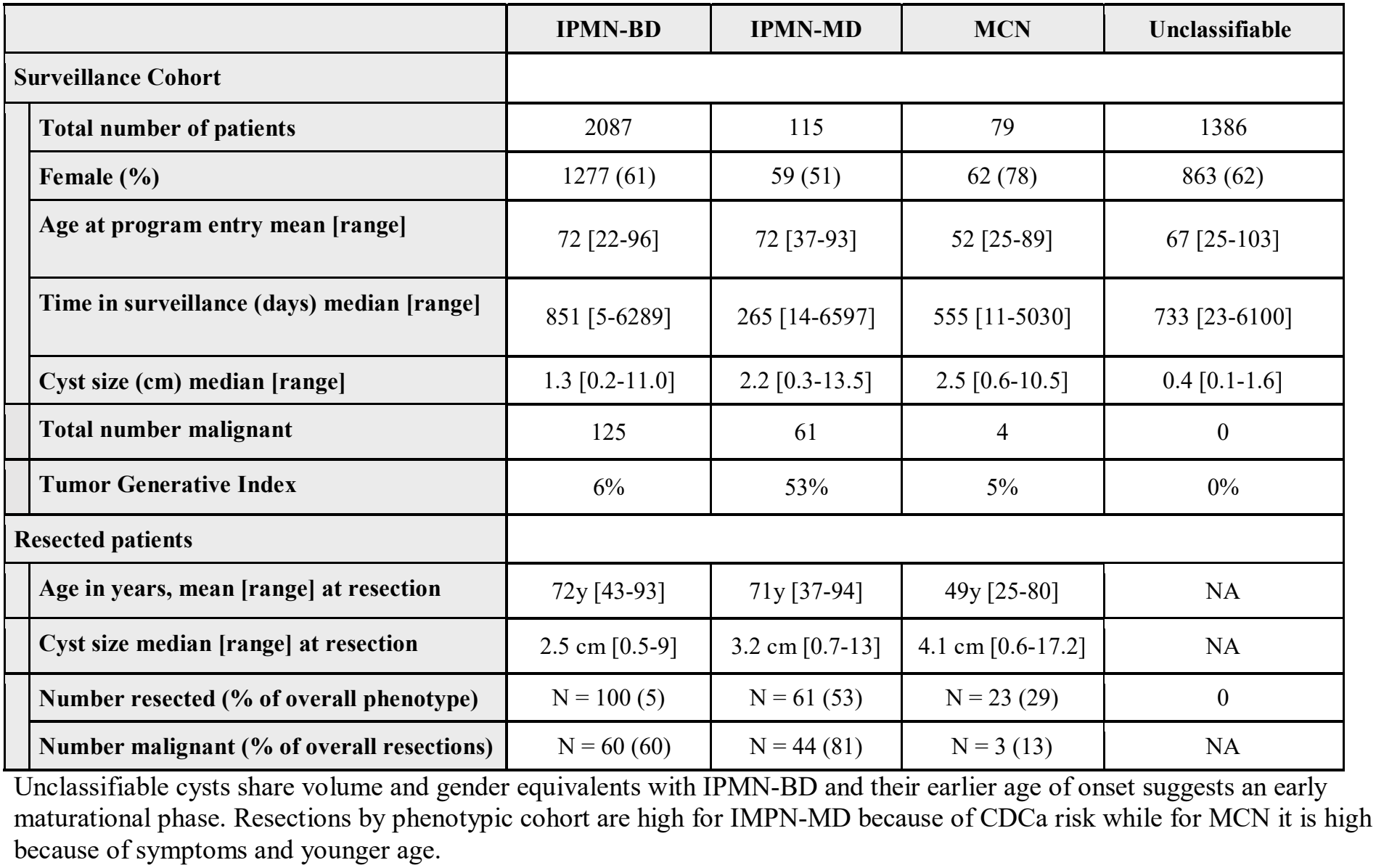
Mucinous cyst profile table.

|  | IPMN-BD | IPMN-MD | MCN | Unclassifiable |
|---|---|---|---|---|
| **Surveillance Cohort** |  |  |  |  |
| **Total number of patients** | 2087 | 115 | 79 | 1386 |
| **Female (%)** | 1277 (61) | 59 (51) | 62 (78) | 863 (62) |
| **Age at program entry mean [range]** | 72 [22-96] | 72 [37-93] | 52 [25-89] | 67 [25-103] |
| **Time in surveillance (days) median [range]** | 851 [5-6289] | 265 [14-6597] | 555 [11-5030] | 733 [23-6100] |
| **Cyst size (cm) median [range]** | 1.3 [0.2-11.0] | 2.2 [0.3-13.5] | 2.5 [0.6-10.5] | 0.4 [0.1-1.6] |
| **Total number malignant** | 125 | 61 | 4 | 0 |
| **Tumor Generative Index** | 6% | 53% | 5% | 0% |
| **Resected patients** |  |  |  |  |
| **Age in years, mean [range] at resection** | 72y [43-93] | 71y [37-94] | 49y [25-80] | NA |
| **Cyst size median [range] at resection** | 2.5 cm [0.5-9] | 3.2 cm [0.7-13] | 4.1 cm [0.6-17.2] | NA |
| **Number resected (% of overall phenotype)** | N = 100 (5) | N = 61 (53) | N = 23 (29) | 0 |
| **Number malignant (% of overall resections)** | N = 60 (60) | N = 44 (81) | N = 3 (13) | NA |

Unclassifiable cysts share volume and gender equivalents with IPMN-BD and their earlier age of onset suggests an early maturational phase. Resections by phenotypic cohort are high for IMPN-MD because of CDCa risk while for MCN it is high because of symptoms and younger age.

### Risk metrics

The frequency of positive risk metric expression within the study cohort included: molecular analysis (n = 47, 10% of tested cysts), solid nodule (n = 213, 6%), Cyst size ≥ 3 cm (n = 255, 7%), pancreatic duct dilation (n = 201, 5%), IPMN-MD (n = 115, 3%), M-PCL-related bile duct obstruction (n = 38, 1%).

### CDCa analysis

The study cohort contained 190 CDCa patients (156 ***ip***CDCa, 34 ***sv***CDCa). CDCa arose from IPMN-BD (n = 125), IPMN-MD (n = 61) and MCN (n = 4). Within the ***sv***CDCa cohort, patients spent a median of 976 days in surveillance and underwent an average of 3 EUS procedures over that time. The inoperability rate in the ***ip***CDCa cohort was 44%, largely from non-resectability (**Figure 1**). Kaplan-Meier survival curves by tumor and nodal status are shown in **Figure 2**. Five-year survival was over 80% for early-stage disease and fell to 50% and 18% for mid-stage and late-stage disease respectively. CDCa-associated nodal positivity by T stage occurred in T1 (4/24, 17%), T2 (13/28, 46%), and T3 (7/20, 35%) of patients (**Figure 2**).

**Fig. 1.**
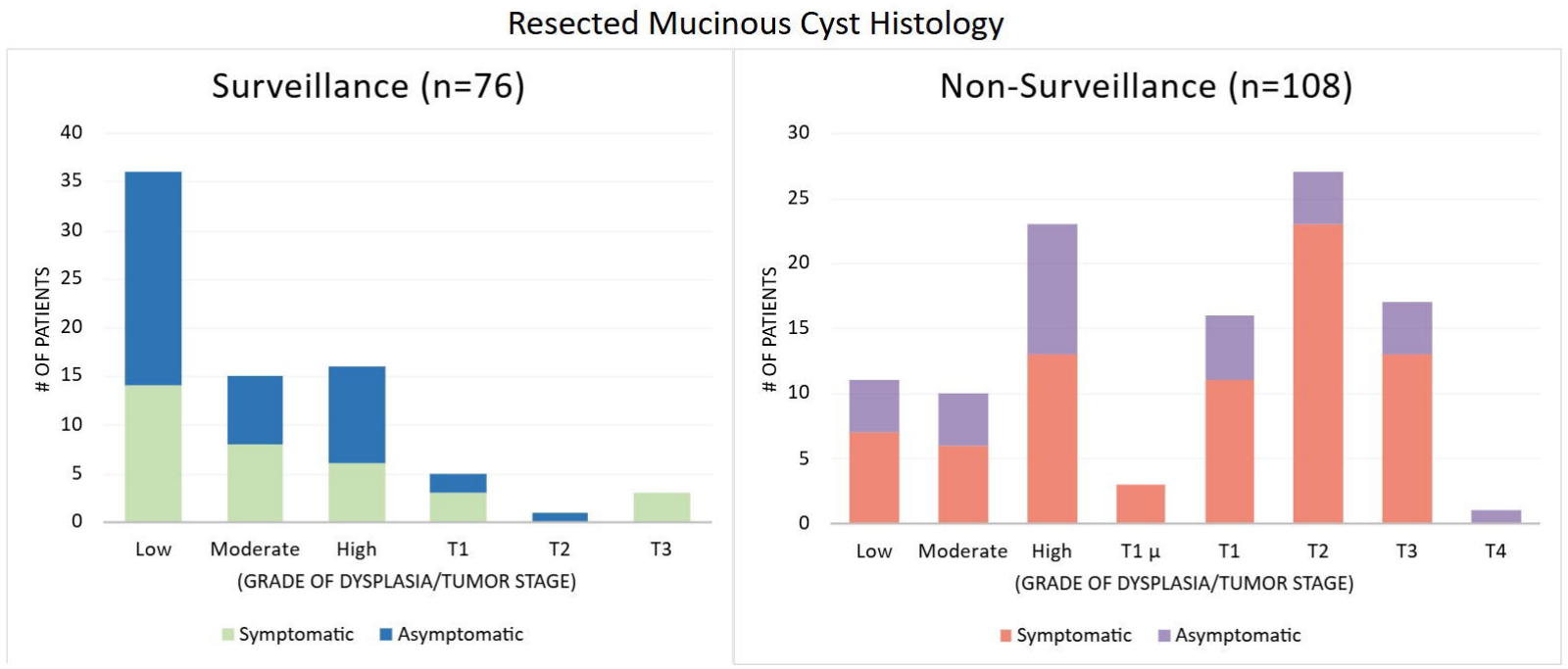

**Fig. 2.**
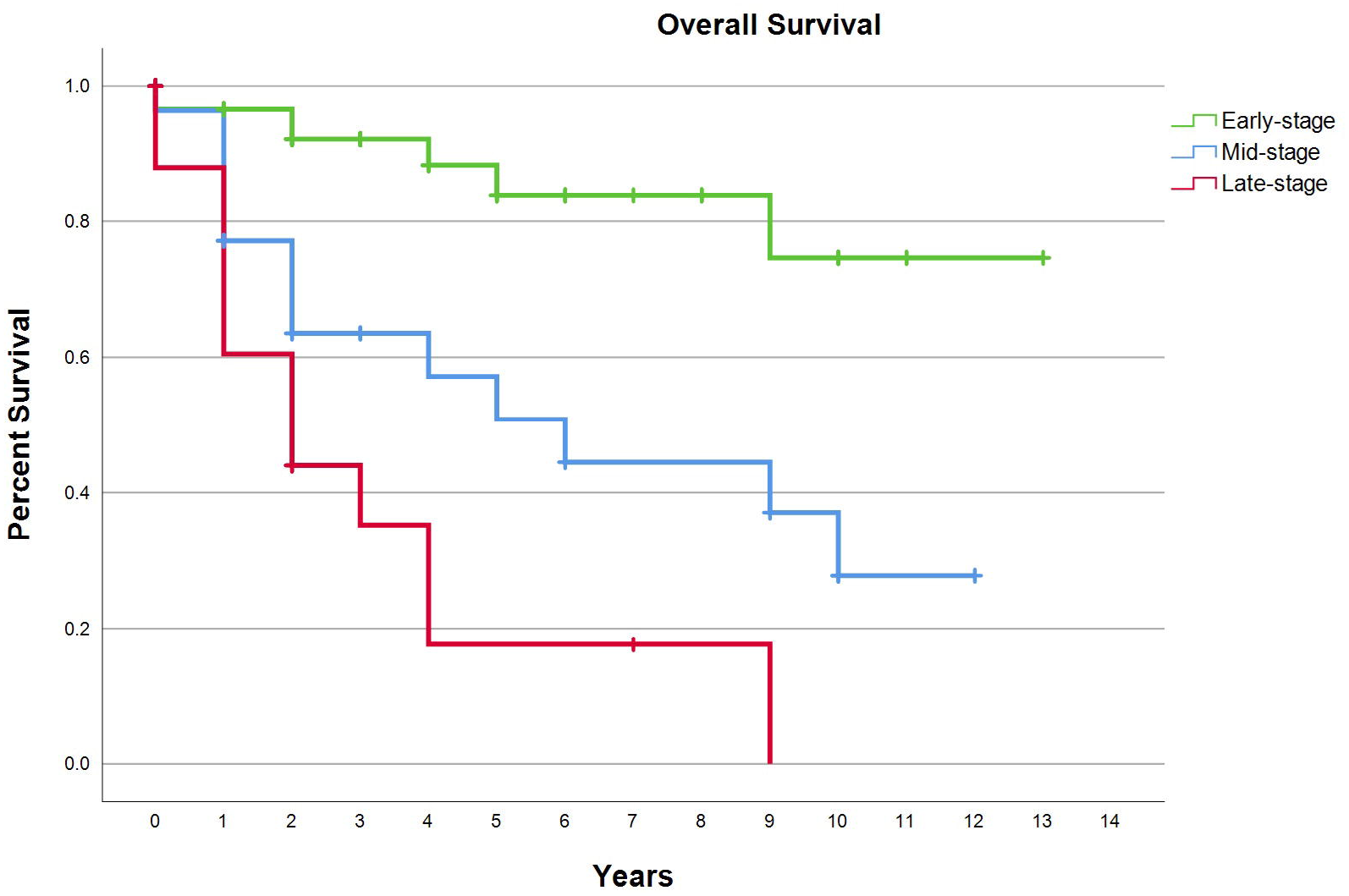

Utilizing our institutional cancer registry data, we determined that 10.7% of all pancreatic adenocarcinoma patients were M-PCL derivatives.

### Surgical outcomes

A total of 184 patients underwent resection over 19 years, 41% from surveillance and 59% at initial presentation. Indications were proven or suspected CDCa (81%) or symptoms (19%). Pancreatectomies included Whipple (42%), distal (49%), total (5%), and central (3%); enucleation comprised the remaining 1%. Surgical mortality was 0%. Six patients required a second resection for progression (n = 3), margin positivity (n = 1), and other reasons (n = 2). Explant pathology revealed 112 of 184 patients (61%) with CDCa, but only 25/112 were identified during surveillance, while 87/112 had CDCa at initial presentation. Seventy-two demonstrated low or moderate grade dysplasia. Resections from non-surveilled patients revealed 81% with CDCa on pathology compared to 33% of patients from the surveillance pathway.

### Risk by cyst phenotype

While most M-PCLs in surveillance were IPMN-BD, the resection percentages by phenotype favored IPMN-MD with a 53% resection frequency, MCN was 29%, and only 5% of the IPMN-BD cohort were resected (**Table 2**). The most striking discordance between preoperative cyst classification and the explant cyst phenotype occurred with IPMN-MD. Of the 61 resected specimens that were confirmed to be IPMN-MD, 27 (44%) were not specifically diagnosed during surveillance. The tumor generative index (TGI) by cyst phenotype was IPMN-MD (53%), IPMN-BD (6%) and MCN (5%).

### Surveillance failures

#### Progression failures

Two CDCa patients from surveillance (6%) were deemed progression failures as their cysts developed malignancy > T1N0 despite compliance with the recommended surveillance studies. They were initially classified as low risk and intermediate risk and were in surveillance for 322 and 330 days respectively.

#### Compliance disruption failures

Nine CDCa patients from surveillance (26%) progressed to malignancy due to compliance failures. Their initial risk categories were high (n = 4), intermediate (n = 4) and low risk (n = 1). The time that elapsed before they reconnected with the program with a diagnosis of CDCa was a median of 1400 days (range 373-2131 days). Three patients (33%) were still operable with T2N0 (n = 1) and T3N0 (n = 2) disease.

### RSS derivation

A total of 3667 samples from 3643 M-PCL patients were included in the model derivation.

#### Model performance

The full six-predictor logistic regression model achieved a cross-validated AUC of 0.956 (95% CI: 0.943–0.967), and the reduced four-predictor model achieved a cross-validated AUC of 0.957 (95% CI: 0.944–0.968), indicating equivalent discriminative performance. DeLong’s test confirmed that the difference in AUC between the two models was not statistically significant (p = 0.893). Comparison of model fit using analysis of deviance and the Akaike Information Criterion also favored the reduced model, supporting parsimony. The ROC curve for the reduced model is shown in **Figure 3**.

**Fig. 3.**
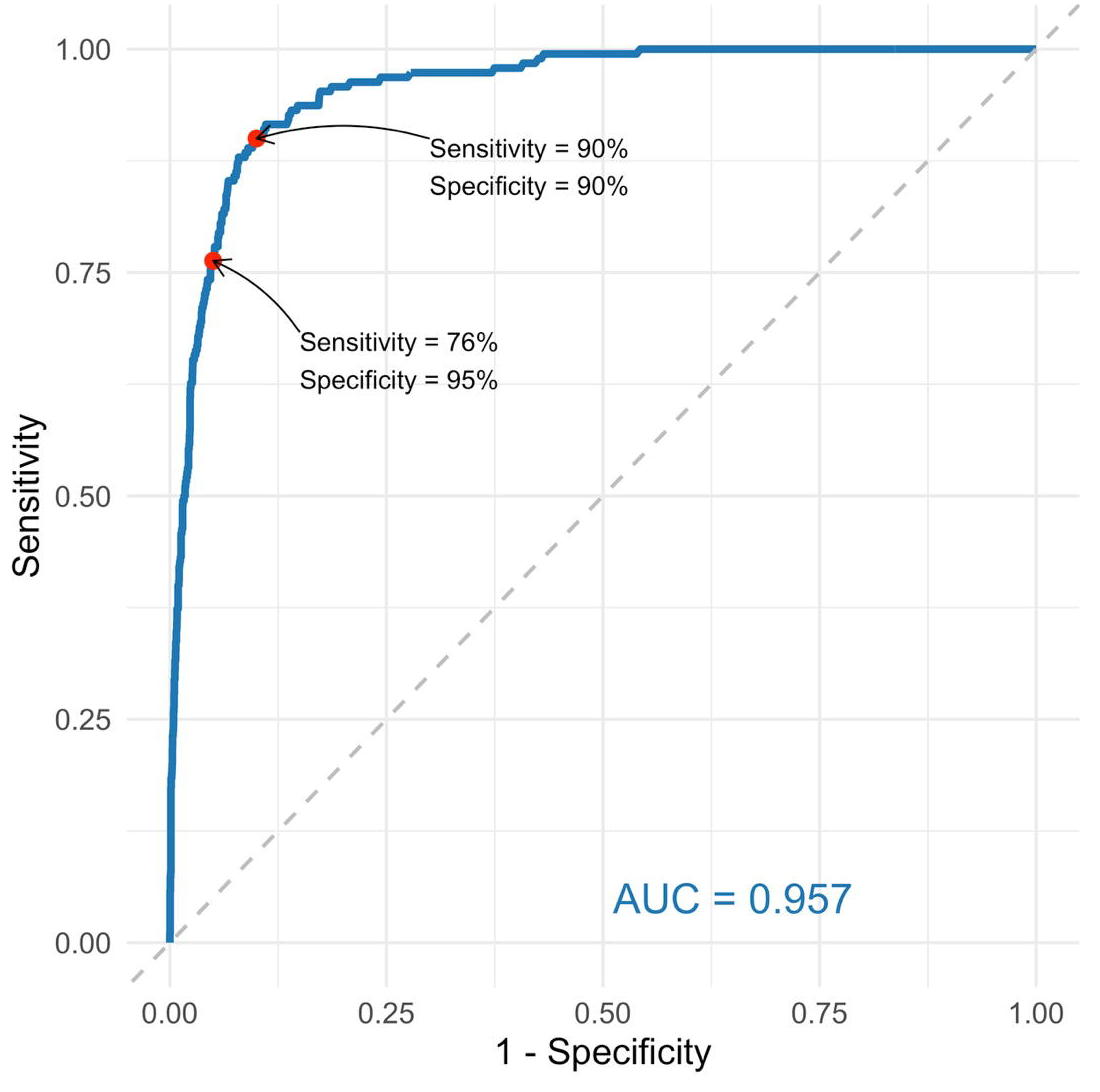

#### Final model

The reduced logistic regression model was selected for parsimony and clinical feasibility. For any new pancreatic cyst evaluated with EUS, it produces an individualized risk score (R) ranging from 0 to 1, representing the predicted probability that the cyst is malignant. The score is based on four EUS-derived predictors: cyst size, main-duct IPMN phenotype, pancreatic duct dilation, and bile duct dilation. Specifically,

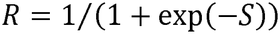

where

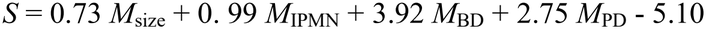

represents a weighted combination of cyst size (*M*_size_, continuous, cm) and binary indicators for main-duct IPMN phenotype (*M*_IPMN_), bile-duct dilatation (*M*_BD_), and pancreatic duct dilatation (*M*_PD_). Predicted risk scores differed markedly between benign and malignant cysts (**Supplemental Figure 2**), with a median of 0.011 (interquartile range 0.009–0.018) for benign lesions and 0.377 (IQR 0.088–0.789) for malignant lesions. Risk-score thresholds corresponding to 90% and 95% specificity were 0.036 and 0.080, respectively, with corresponding sensitivities of 90% and 76%.

## Discussion

Due to an aging population and increased recognition, mucinous pancreatic cysts have become a population health problem. Like other published studies, our experience confirms an increased incidence and prevalence of mucinous cysts.^24^

Despite this, there has yet to be a single highly accurate risk metric that is accessible through blood or cyst fluid which can predict a patient’s risk of cyst-derived malignancy. This leaves clinicians relying on a mosaic of clinical, radiologic and biochemical risk features to guide patient management. This is the point where surgical decision making is simultaneously data-driven yet intuitive. It is also in stark contrast to the diagnosis and management of other solid organ malignancies which offer tissue diagnosis prior to determining clinical management. Existing society practice guidelines focus on recommendations for screening intervals and broad principles of management, leaving the field without an objective measure to predict malignancy risk versus benign disease. Our risk probability score aims to address this gap and help facilitate early recognition of oncogenesis at a stage amenable to surgical cure.

Adding complexity to the decision-making process is the high prevalence of pancreatic cysts in geriatric patients for whom surgery carries a higher risk of morbidity and mortality.

Cyst-derived cancer occurred in 8% of our largely geriatric mucinous cohort. This prompts a great degree of consideration when balancing the potential risks and benefits of an operation and underscores the necessity of having a reliable risk assessment metric to inform clinical decision making.

Historical models emphasize cancer risk sensitivity, often at the expense of specificity.^4,27^ Our own experience reflects this as well. From the entire M-PCL surgical cohort, 39% of surgical explants demonstrated low to moderate grade dysplasia. This remained the case even when highly symptomatic patients with benign pathology were censored from the cancer-risk analysis. Patient and physician fears of missing CDCa must be balanced when recommending an operative risk/recovery that can be challenging.

In developing our new risk stratification score, we used logistic regression analysis to identify the four most predictive risk metrics for CDCa (cyst size, pancreatic duct dilation, bile duct dilation, IPMN-MD) in a weighted formula used to compute the probability of malignancy of a M-PCL. Achieving an AUC value of 0.96, the model proved extremely accurate at assessing mucinous cysts for malignancy. This improved upon our original risk stratification protocol which relied on assigning parity to all risk metrics and resulted in a lack of specificity. Our new model allows us to toggle along the accuracy curve (**Figure 3**) to balance its sensitivity and specificity. Optimizing specificity is especially important to both surgeons and patients alike when facing high risk pancreatic surgeries with difficult recoveries. We still recommend continuation of surveillance based upon individual risk metrics even for the lowest risk and unclassifiable cyst patients but feel that the new derivation can be combined with specific individual patient considerations to facilitate decision making.

Analysis of our patient cohort yielded several additional novel observations which offer further insight into the nuances of pancreatic cyst surveillance and management. First, we noted that cyst-derived pancreatic cancer appears to have a more favorable tumor biology. Lymph node negative pathology was much more common in CDCa patients, even with T2/T3 lesions, and survival more favorable when compared with PDAC, as outlined in **Figure 2**. This difference in oncologic behavior can offer a forgiving cushion for early CDCa and allows further confirmation of progression through tightened surveillance prior to major surgical commitment. While this favorable tumor biology of cyst-derived pancreatic cancer has been previously described in the literature, other single-center series have observed rates of nodal metastases as high as 22% and some have suggested that the higher survival is related to earlier resection due to surveillance.^25,26,30–33^ This highlights a need for further study into the true histopathologic behavior of cyst-derived pancreatic cancer.

Secondly, we identified a specific phenotypic discordance between preoperative labeling of phenotypic cyst type and final pathologic confirmation. In our data, the distinct cyst phenotypes demonstrated numeric, demographic and oncogenic differences, thus making accurate cyst type identification crucial during surveillance. Particularly troubling is that 44% of patients with main duct involvement on final pathology were classified as IPMN-BD preoperatively. Historic series have documented a similar difficulty with preoperative classification of cysts, finding main duct involvement in 20-29% of lesions which were labeled as IPMN-BD on preoperative imaging.^28,29^ Under-recognition of IPMN-MD in the surveillance cohort is concerning given the high TGI of this phenotype.

Finally, we noted that unclassifiable cysts made up a large subset in our cohort. Our longitudinal observation of these cysts and comparison of other anatomic and demographic findings of these patients leads us to believe that these may be primordial IPMN-BDs. These findings emphasize the necessity of long-term longitudinal follow-up in these patients and caution against prematurely designating them as safe based on size alone.

Although a structured surveillance plan works to improve early cancer recognition, the unfortunate truth is that CDCa was discovered five times more often at initial presentation than through surveillance, resulting in advanced tumors and lower operability. Even active patients within the program can become non-compliant after protracted follow-up resulting in late stage CDCa. This underscores the need to disseminate validated guidelines for physicians and patients to increase surveillance practice based on improved understanding of the natural history of the disease.

## Supporting information

Supplemental Figure 1: Surveillance patient activity

Supplemental Figure 2: Risk score comparison

## Data Availability

The data that support the findings of this study are not available, as research participants did not provide written consent for their data to be shared publicly or with third parties.

## Notes

### Competing Interest Statement

The authors have declared no competing interest.

### Author Declarations

IRB of Hoag Memorial Hospital Presbyterian gave ethical approval for this work.

