## Supplementary figures and images for "Mucinous Pancreatic Cysts: A Single-Center Experience Epidemiology, Surveillance, Risk Formulation, Surgery"

### Supplemental Figure 1: Surveillance patient activity

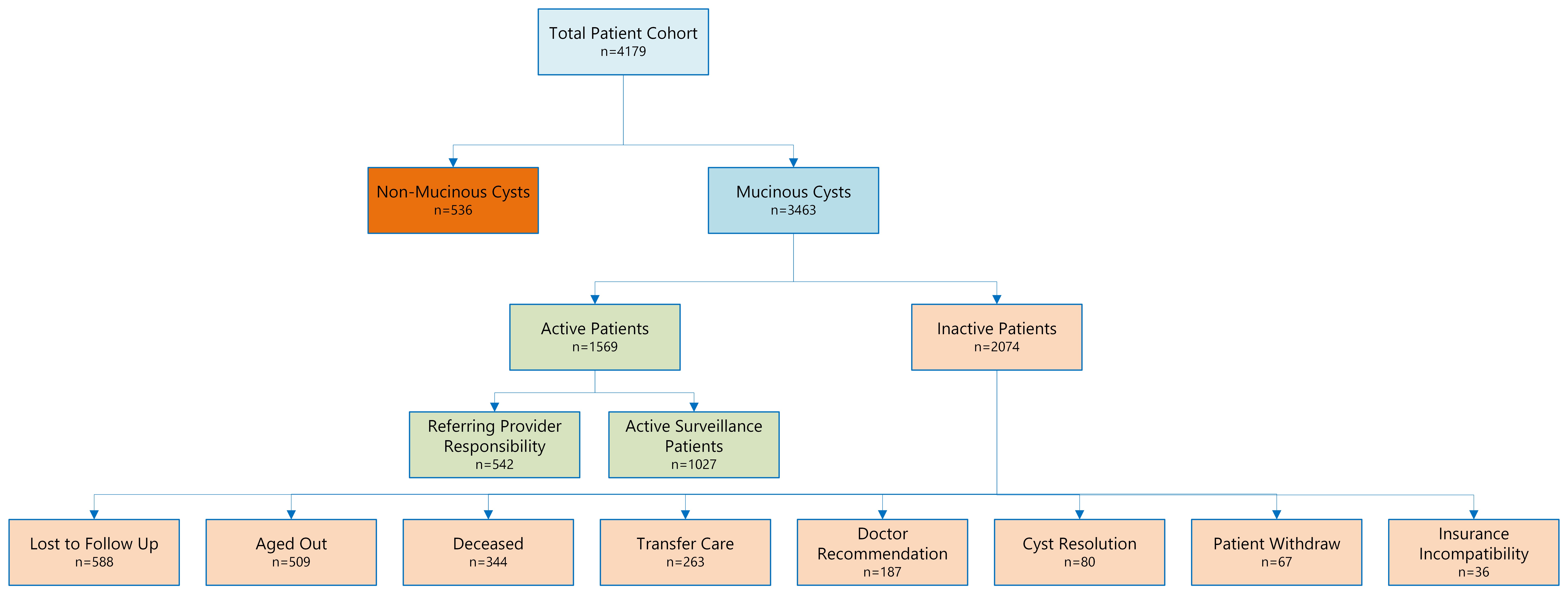

### Supplemental Figure 2: Risk score comparison

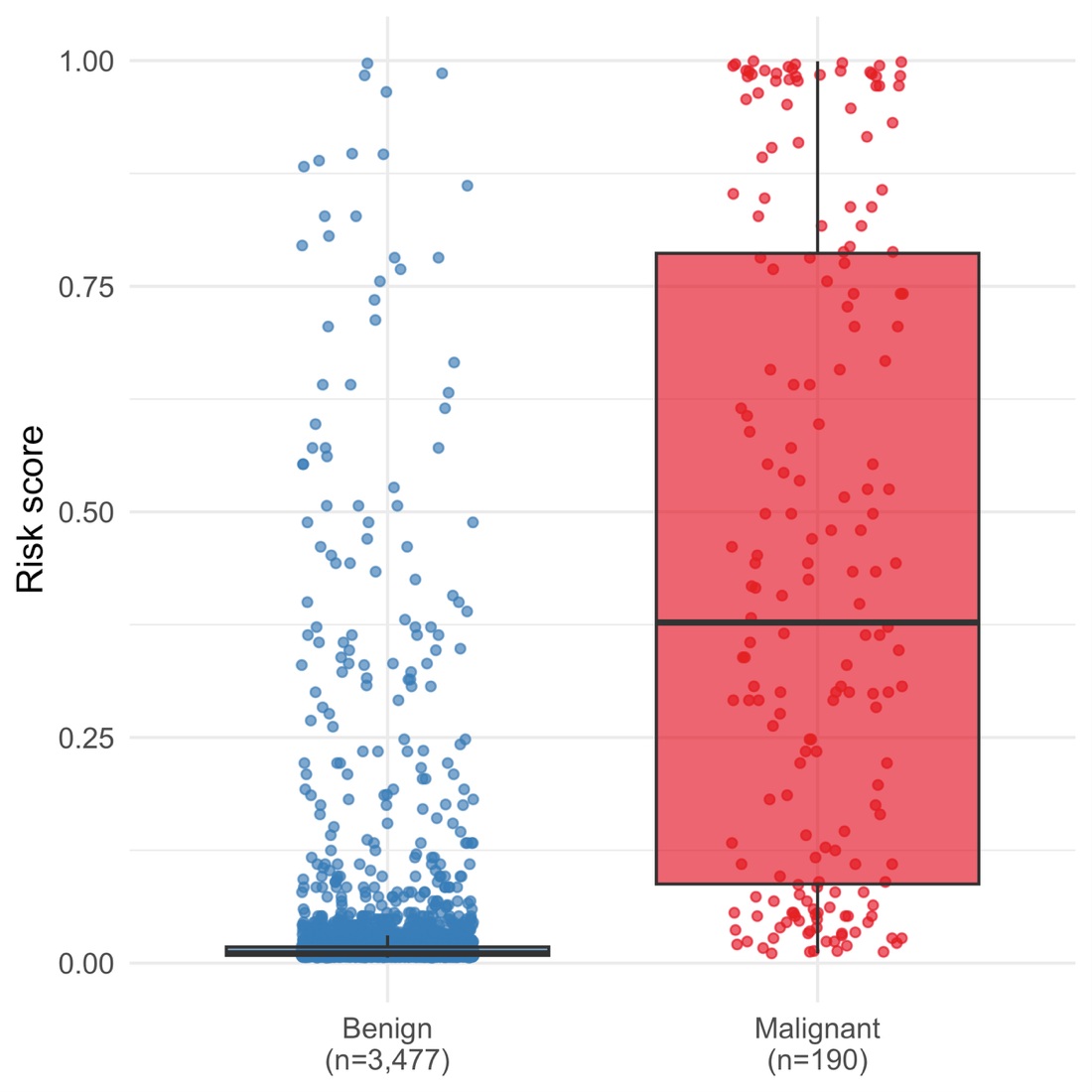
